# Appropriate sampling and long follow-up are required to rigorously evaluate longevity of humoral memory after vaccination

**DOI:** 10.1101/2023.06.28.23291950

**Authors:** Vitaly V. Ganusov

## Abstract

One of the goals of vaccination is to induce long-term immunity against the infection and/or disease. However, evaluating the duration of protection following vaccination often requires long-term follow-ups that can conflict with the desire to rapidly publish results. Arunachalam et al. JCI 2023 followed individuals receiving third or fourth dose of mRNA COVID19 vaccines for up to 6 months and in finding that the levels of SARS-CoV2-specific antibodies (Abs) declined with similar rates for the two groups came to the conclusion that additional boosting is unnecessary to prolong immunity to SARS-CoV-2. However, this may be premature conclusion to make. Accordingly, we demonstrate that measuring Ab levels at 3 time points and only for a short (up to 6 month) duration does not allow to accurately and rigorously evaluate the long-term half-life of vaccine-induced Abs. By using the data from a cohort of blood donors followed for several years, we show that after re-vaccination with vaccinia virus (VV), VV-specific Abs decay bi-phasically and even the late decay rate exceeds the true slow loss rate of humoral memory observed years prior to the boosting. We argue that mathematical modeling should be used to better optimize sampling schedules to provide more reliable advice about the duration of humoral immunity after repeated vaccinations.

---

“To consult the statistician after an experiment is finished is often merely to ask him to conduct a post mortem examination. He can perhaps say what the experiment died of. “ Ronald Fisher.

## Results and discussion

The holy grail for vaccinologists is to induce long term effective immunity against infections, such as against the SARS-CoV-2, the cause of the recent pandemic^1^. It is well known that specific antibodies (**Abs**) to the spike protein of SARS-CoV-2 account for disease protection^2^, and it is of great interest to discover how long such antibodies persist after infection or various forms and regimens of vaccinations^3,4^. In one such study, Arunachalam *et al*. ^4^, compared the durability of neutralizing antibody (**nAb**) levels in persons that received either 3 or 4 vaccinations with Pfizer or Moderna mRNA vaccines. By using a simple linear regression, Arunachalam *et al*. ^4^ showed that the decay of Abs between 1 and 6 months was similar in individuals receiving 3rd or 4th vaccination allowing them to conclude that “The durability of serum antibody responses improves only marginally following booster immunizations with the Pfizer-BioNTech or Moderna mRNA vaccines.” inferring that extra boosters are unnecessary to confer long-term immunity.

From multiple studies in mice and humans it is clear that the kinetics of Ab decay are generally more complex than a simple exponentially decaying function would suggest^5–7^. Specifically, after the peak immune response, Ab titers initially decline rapidly and approach their long-term decay kinetics over months. To investigate how the dynamics of Ab titers after revaccination relates to their long-term maintenance, we re-analyzed unique data from a cohort of long-term blood donors that recorded Ab titers to multiple vaccines over a long time period^8^. For this analysis, we selected 4 individuals illustrated in Figure 2 of Amanna *et al*. ^8^ that were followed for 20+ years and subsequently been revaccinated with vaccinia virus (**Figure 1** and **Supplemental Figure S1**). The kinetics of Ab decay prior to revaccination showed remarkable stability of VV-specific Abs with half-life times of 31-91 years (**Figure 1**A). By fitting a modification of the mathematical model of Ab response reported previously^7^ to the data on Ab expansion and contraction after revaccination, we found much smaller half-life times of VV-specific Abs 1.4-2.6 years as compared to the half-life times prior to revaccination (except of one volunteer (ID=514) showing no long-term decay after revaccination, **Figure 1**B). Given that these individuals were followed for nearly 2 years after VV revaccination their VV-specific Abs levels still decayed more rapidly than during years prior to boosting. This analysis indicates that long follow up studies are required before any conclusions can be made about the true longevity of vaccine-induced Abs. Additional analysis of the kinetics of Ab decay after the peak following revaccination with VV or tetanus vaccine suggested that these decays are not well described by a simple exponential curve indicating that different sub-populations of antibody-secreting cells (**ASCs**) with discordant lifespans may be present (**Supplemental Figure S1**). Interestingly, the long-term loss of tetanus-specific Abs occurred at a slower rate than the loss of VV-specific Abs, which may be surprising since VV is a live vaccine and tetanus vaccine is a protein-based vaccine. The difference here likely lies in the different times of follow up allowing more rigorous estimates of the half-life time of tetanus-specific Abs (**Supplemental Figure S1**).

**Figure 1:**
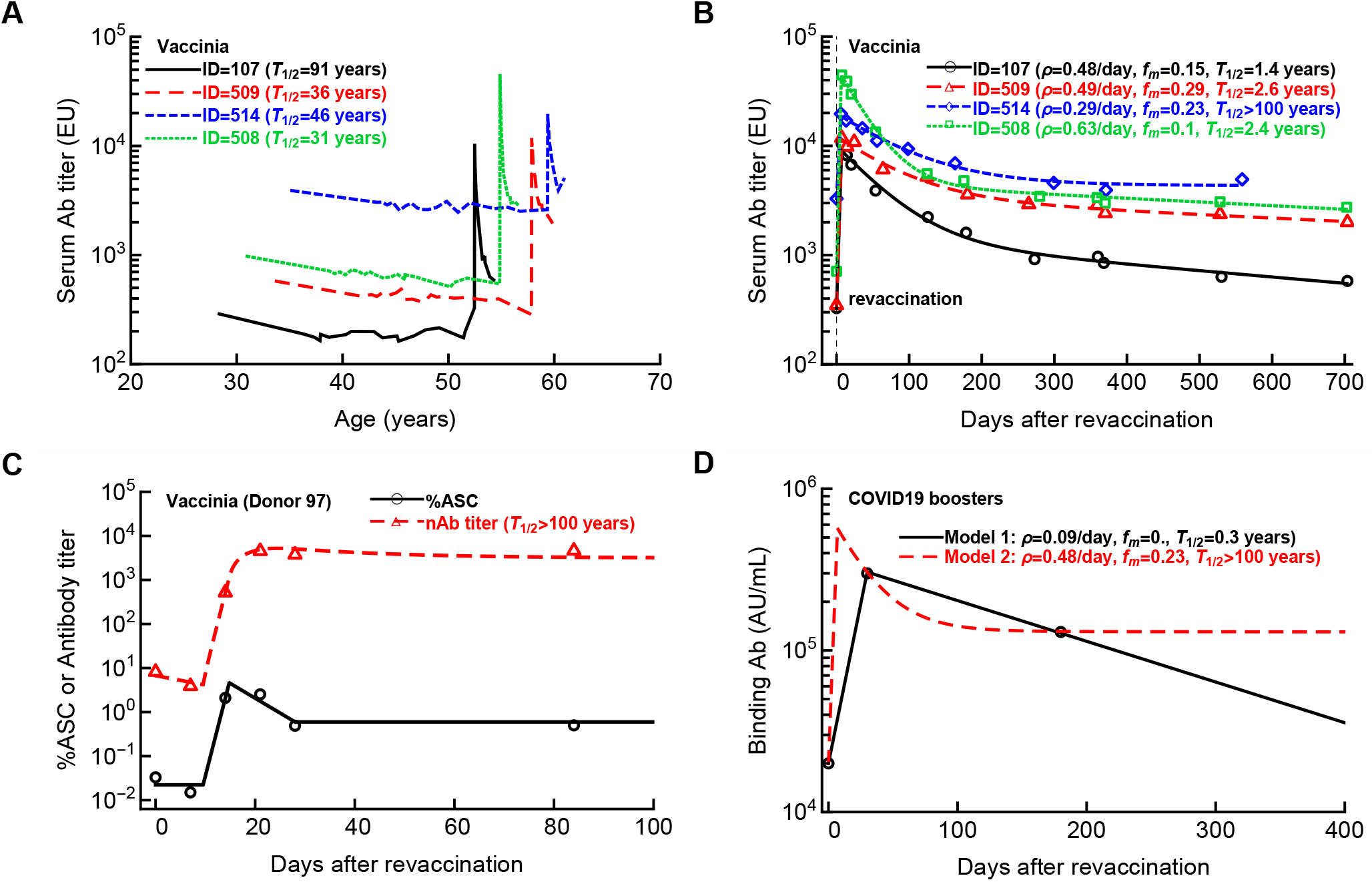
Stable levels of antibodies are reached only months after revaccination with vaccinia virus. **A**: We analyzed kinetics of Ab titers in four long-term blood donors from subjects 1-4 shown in Figure 2 of Amanna *et al.*^8^). These individuals were followed up for 20+ years during which they had been revaccinated with vaccinia virus. **B**: We fitted a mathematical model of humoral immune response (**eqn. (S.1)**)) to subsets of the data that include revaccination and estimated the rate Ab expansion (*ρ*), the proportion of Ab conversion into long-lived population (*f*_*m*_), and the half-life of the humoral immunity (*T*_1/2_, panel B and see **Supplemental Table S1** for estimated model parameters). **C**: Kinetics of Ab response following VV revaccination in one volunteer (Donor 97) suggests infinite half-life of the long-term memory (see Main text for best fit parameters)^7^. **D**: Sparse measurements of Ab titers after vaccination allow for alternative mathematical models (**eqn. (S.1)** with different parameter sets) with drastically different predicted longevities of Abs. Here the markers are average Ab titers from Figure 1Ciii of Arunachalam *et al.*^4^, and lines are predictions of two alternative mathematical models with different assumed sub-populations of ASCs.

In our recent work, we formulated mathematical models aimed to describe kinetics of ASCs and Ab titer following vaccination of humans^7^. The example of the immune response following VV immunization showed that to capture Ab loss, one needs to measure Ab titers frequently around the peak of the immune response (**Figure 1**C; VV-specific response for Donor 97 in the model given in **eqns. (S.2)–(S.3)** are *P*_0_ = 0.022, *T*_on_= 9.5 day, *ρ* = 1.02 day, *T*_off_ = 14.79 day, *δ*_*A*_ = 0.15/*day, T*_mem_ = 28 day, *A*_0_ = 6.7, *p* = 261.3/*day*) ^7^.

We then used the model that described well the Ab response to VV (**eqn. (S.1)**) and selected parameters that would allow matching the average Ab titers measured at three time points chosen in the report of Arunachalam *et al*. ^4^. Importantly, we could choose two extreme sets of parameters that predict either relatively short-lived Ab response with half-life *T*_1/2_ = 0.3 years (model parameters: *A*_0_ = 20000, *ρ* = 0.091/*day, T*_off_ = 30 day, *f*_*m*_ = 0, *δ*_1_ = 0.0058/*day, δ*_2_ = 0.13/*day*) or extremely long Ab response (*T*_1/2_ *>*100 years, model parameters: *A*_0_ = 20000, *ρ* = 0.48/day, *T*_off_ = 7 day, *f*_*m*_ = 0.23, *δ*_1_ = 0.04/*day, δ*_2_ = 0.000013/day, **Figure 1**). These results further demonstrate that measuring Ab titers at only 3 time sparsely spaced points does not allow to rigorously evaluate the duration of humoral memory following revaccination.

What are the possible solutions? Obviously, following revaccinated individuals for longer times may provide some useful information, but inapparent boostings of immunity following reinfection from currently circulating SARS-CoV-2 cannot be excluded. A better approach would be to improve the experimental design by using mathematical modeling-assisted power analyses that should indicate the most informative time points that samples should be collected at to provide the most reliable estimate of the parameter of immunity that is being advocated such as the Ab half-life times^9,10^. Following Ab response longitudinally in individual volunteers may also allow to use a more powerful approach of mixed effect modeling that may help to better define average and variability in Ab half-life times^11^. Ultimately, better collaborations between experimentalists and mathematical modelers may help to design more reliable experiments that provide more rigorous estimates of the longevity of humoral immunity afforded by vaccination, and (to paraphrase Ronald Fisher) “not let postmortem analysis to identify reasons experiment died of”.

## Supporting information

Data on Ab dynamics following vaccinia virus or tetanus revacciation

## Data Availability

as supplement to the paper

## Acknowledgement

We would like to thank Mark Slifka and Ian Amanna for providing experimental data for this analysis. This was work supposed by the NIH grant (R01AI158963) to VVG.

## Abbreviations

VV: vaccinia virus
EU: ELISA units
NLS: nonlinear least squares
ASCs: antibody-secreting cells
NLME: nonlinear mixed effects.

## Supplemental information

### Materials and Methods

#### Experimental data

Experimental data on kinetics of Ab titers in blood donor volunteers have been provided by Drs. Mark Slifka and Ian Amanna and are from their previous publication^8^. Here we specifically analyzed the data on the kinetics of VV and tetanus-specific Abs from four subjects shown in Figure 2 of Amanna *et al*. ^8^. We focused specifically on VV and tetanus-specific Abs because revaccination with these antigens induced robust recall responses in all individuals. To fit mathematical models to the Ab kinetics after revaccination we only included the first measurement before the peak of response and all the available data after revaccination. More thorough analysis of the recall responses in other subjects and to other vaccines will be provided elsewhere. The analyzed data (VV- and tetanus-specific Ab titers) are provided with the submission. Data on the dynamics ASCs and Ab titers following boost immunization with vaccinia virus (Donor 97) was published previously^7^. As far as we are aware the listed donor IDs are not known to anyone outside the research group that performed these measurements.

#### Mathematical model for the kinetics of humoral immune response

In experiments involving immunization, it is common to only measure the Ab titers (either antigen-binding titers or netraulizating titers). To describe the kinetics of Ab response for those data we adapt a simple *T*_on_ − *T*_off_ model proposed to describe kinetics of the T cell response to viruses^7,12,13^. In this model, immune response gets activated at *T*_on_ time and grows exponentially at a per capita rate *ρ*. The immune response peaks at time *T*_off_ and then decays at two rates, *δ*_1_ and *δ*_2_, that biologically corresponds to ASCs with different longevity (given at frequency 1 *− f*_*m*_ and *f*_*m*_, respectively):

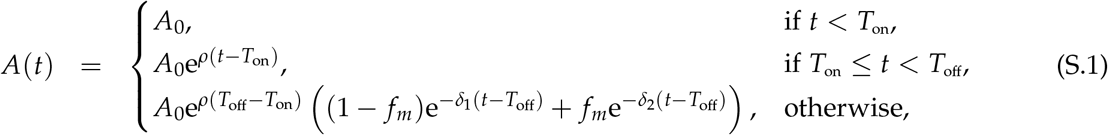

where *A*_0_ is the initial Ab titer (prior to boosting), *T*_on_ is the time of the boosting (typically, *T*_on_ = 0), *ρ* is the expansion rate of the Ab response, *T*_off_ is the time of the peak of the immune response, *δ*_1_ and *δ*_2_ are the rates of Ab decay in the short- and long-lived plasma cells, and *f*_*m*_ is the proportion of the long-lived plasma cells in the total ASC populations. Note that the half-life of humoral memory is determined by the lowest of the two decay rates *δ*_1_ and *δ*_2_, *T*_1/2_ = log(2)/ min(*δ*_1_, *δ*_2_).

To describe the dynamics of both ASC and Ab titer response following revaccination we use a model proposed previously^7^. In model, ASC number (*P*) expands exponentially after a delay *T*_on_, peaks at *T*_off_, and then ASCs die and convert to long-lived plasma cell phenotype at a rate *δ*_*A*_. At *T*_mem_ long-lived plasma cells are formed; they decay at rate *δ*_*M*_. ASCs produce Abs at a rate *p* and Abs decay at a rate *δ*_*a*_:

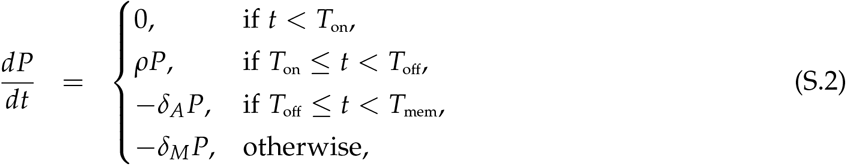

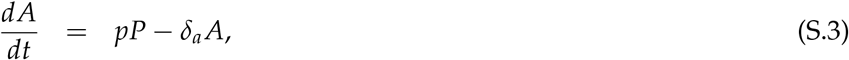

where *p* is the per capita rate of production of antibodies by virus-specific plasma cells, and *δ*_*a*_ is the natural decay rate of antibodies assumed to be *δ*_*a*_ = 0.0495 day^*−*1^ corresponding to a half-life of 2 weeks^14,15^. In our example, *δ*_*M*_ = 0 as this provided the best fit of the model to data^7^.

To describe the kinetics of Ab decay after the peak following VV or tetanus revaccination we used several alternative mathematical models that included a different number *n* of subpopulations of ASCs with different lifespans:

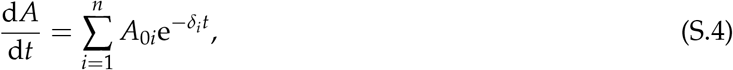

where *A* is the Ab titer from all sub-populations, *A*_0*i*_ is the size of the *i*^*th*^ sub-population and *δ*_*i*_ is the decay rate of ASCs in the *i*^*th*^ sub-population. The ultimate half-life of Abs is determined by the slowest decay rate *δ* = min(*δ*_*i*_) with the corresponding half-life time *T*_1/2_ = ln(2)/*δ*. To determine how many sub-populations are needed to best describe the kinetics of Ab loss after the peak we vary *n* = 1 … 3 and compare the quality of the model fits using F-test for nested models^16^.

#### Statistics

The models were fitted to data using nonlinear least squares (NLS) by log-transforming the model predictions and the data. We used routine `FindMinimum` in Mathematica for NLS.

## ADDITIONAL FIGURES AND TABLES

**Supplemental Figure S1:**
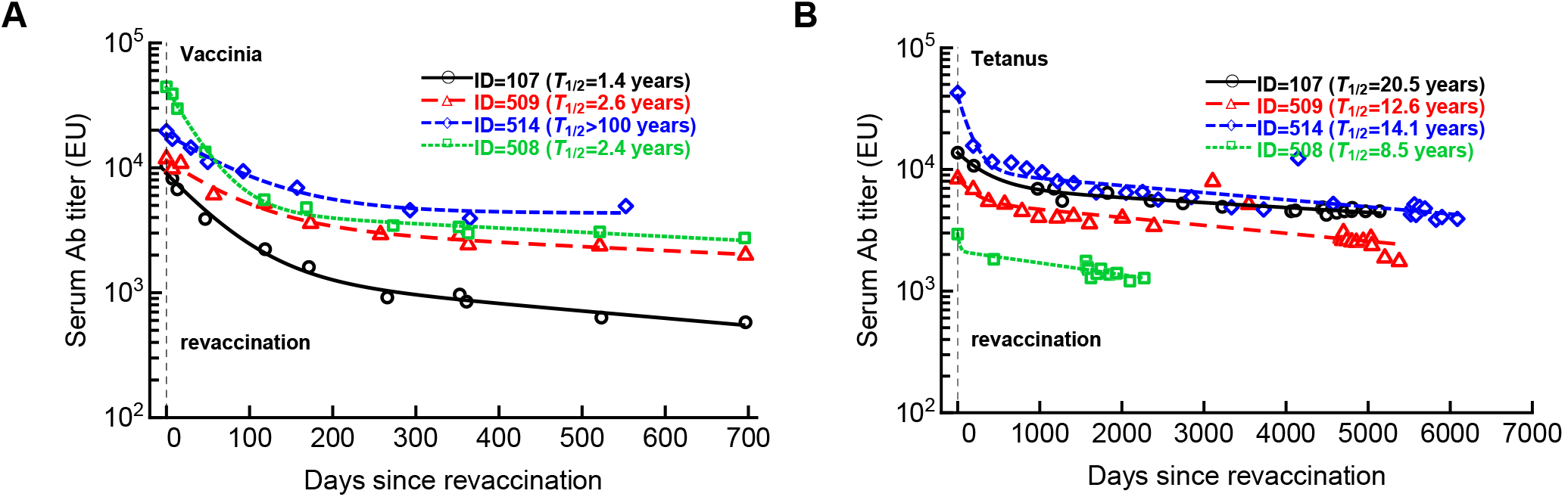
Approach to stable Ab levels takes months after revaccination with vaccinia virus or tetanus vaccine. We fitted several alternative mathematical models for Ab decay (**eqn. (S.4)**) to data by varying the number of sub-populations for four volunteers revaccinated with VV (**A**) or tetanus (**B**) vaccines. In the analysis we included only the data from the peak Ab response at the first instance after an apparent revaccination (noted by the horizontal dashed line). In both cases the models with 2 subpopulations fitted the data better than the model with a single sub-population but a larger model (*n* = 3) did not improve the model fit quality but did provide a longer half-life times for persisting Abs. For each fit we provide the estimated half-life time (*T*_1/2_)based on the slowest decay rate. Parameters of the best fit models are show in **Supplemental Table S2**. Note the different scales of the follow up for boosting with two different vaccines.

**Supplemental Table S1:** Estimates of parameters of the Ab expansion and decay kinetics during revaccination with VV. We fitted mathematical models (**eqn. (S.1)**) to the data on Ab expansion and decay kinetics in four volunteers following revaccination with VV (**Figure 1**B).

| ID | vaccine | $A_0$ | $\rho$, 1/day | $T_{\text{off}}$, day | $f_m$ | $\delta_1$, 1/day | $\delta_2$, 1/day |
| --- | --- | --- | --- | --- | --- | --- | --- |
| 107 | VV | 327.2 | 0.48 | 6.94 | 0.15 | 0.018 | 0.00133 |
| 509 | VV | 348.8 | 0.49 | 7.09 | 0.29 | 0.014 | 0.00072 |
| 514 | VV | 3284.4 | 0.29 | 6.07 | 0.23 | 0.012 | 0 |
| 508 | VV | 710.7 | 0.63 | 6.5 | 0.10 | 0.029 | 0.00078 |

**Supplemental Table S2:** Estimates of parameters of the Ab decay mathematical model fitted to data on Ab kinetics in four volunteers vaccinated with VV or tetanus vaccine. We fitted several mathematical models (**eqn. (S.4)**) to the data on Ab decay in four volunteers following revaccination with VV or tetanus vaccine (**Supplemental Figure S1**).

| ID | vaccine | $A_{01}$ | $A_{02}$ | $A_{03}$ | $\delta_1$, 1/day | $\delta_2$, 1/day | $\delta_3$, 1/day | $T_{1/2}$, year |
| --- | --- | --- | --- | --- | --- | --- | --- | --- |
| 107 | VV | 7619.0 | 1387.9 | 0 | 0.0181 | 0.0013 | 0 | 1.45 |
| 509 | VV | 8123.8 | 3340.9 | 0 | 0.0138 | 0.0007 | 0 | 2.68 |
| 514 | VV | 14365.2 | 4335.8 | 0 | 0.0121 | 0 | 0 | > 100 |
| 508 | VV | 39419.9 | 4502.3 | 0 | 0.0293 | 0.0008 | 0 | 2.47 |
| 107 | Tetanus | 7038.6 | 6675.2 | 0 | 0.00009 | 0.00285 | 0 | 20.83 |
| 509 | Tetanus | 3096.0 | 5444.1 | 0 | 0.00524 | 0.00015 | 0 | 12.82 |
| 514 | Tetanus | 30789.1 | 9646.3 | 0 | 0.00689 | 0.00013 | 0 | 14.32 |
| 508 | Tetanus | 816.4 | 2127.5 | 0 | 0.04081 | 0.00022 | 0 | 8.66 |

